# Anthropometric measurements in children with ASD point to genomic imprinting imbalance

**DOI:** 10.1101/2020.04.07.20043224

**Authors:** Alexandru-Ștefan Niculae

## Abstract

Autism Spectrum Disorder (ASD) is a large set of neurodevelopmental disorders of complex aetiology. A mix of genetic and environmental factors are likely to cause ASD. Genetic risk for autism comes from common genetic variation.

Genomic imprinting refers to genes that have different expression patterns according to the parent of origin – being silenced when imprinted. Paternally active genes increase resource extraction from the mother and reduce resource burden on the father.

Children with ASD show consistent overgrowth during their first 1-2 years of life. Recently, it has been shown that children with higher birth weight and length have an increased risk of developing ASD. This overgrowth and apparent larger birth weight and length are consistent with the notion that a paternally biased genome might underlie the risk for ASD.

The study compared height, weight, head circumference and thoracic circumference for age-matched (ages 4-8 years old) male children with ASD (n=30) with neurotypical children (n=33).

No clinically significant differences were found among the two groups.

After weaning, relative paternal contribution to a child’s somatic development would increase, thus one would expect paternally active genes to start changing the child’s behaviour, so as to make the child less demanding of resources (overall, and thus also on the father), with a counterweight represented by maternally active genes. A relative overabundance of paternally active genes would explain the data presented here, that shows children with ASD being no different from controls. Given the fact presented by other studies, that children with ASD seem to get a head start in growth, the lack of differences found in this 4-8 years old group indicates that children with ASD might actually fall behind in somatic growth, or at least stagnate by middle childhood.

## INTRODUCTION

Autism Spectrum Disorder (ASD) affects social interactions, language and communications with peers and other members of the group and overall behaviour.

Genetic factors account for most of the risk underlying the development of ASD^1^ and genetic risk for autism comes from common genetic variation^2^.

Genomic imprinting refers to epigenetic changes that regulate the gene’s expression. Genes that are imprinted are not transcribed. The imprint is passed down from parents to offspring, and an offspring’s somatic cells will carry the imprints inherited from the parents, with corresponding gene expression patterns. Current understanding of the emergence and function of imprinted genes is formalised as the kinship theory of genomic imprinting ^3^.

Genes that have different expression patterns according to the parent of origin (i.e. inactive when imprinted) lead to (relatively) increased resource extraction from the opposite parent. Paternally active genes increase resource extraction from the mother and reduce resource burden on the father ^3,4^, both during foetal development and early infancy, but also extending after weaning into later childhood ^3^.

Somatic growth patterns in children with ASD suggest a relative overabundance of paternally active genes. Studies have analysed anthropometric data in children since birth or early infancy. As children aged, those who went on to develop ASD were shown to have had higher growth rates compared to peers. These studies shown that: (i) head circumference increased faster in the first year of life in children who later develop ASD ^5^, (ii) both head circumference and corresponding overall weight and length are larger in children who later go on to develop ASD ^6^, (iii) and most recently, faster weight gained during infancy increases the risk for later developing ASD by up to three times ^7^.

There is also evidence from a large retrospective Danish cohort study that higher than average birth weights and lengths also carry a significantly higher risk for developing ASD later in life ^8^.

This summarized evidence, that has already been published, supports the notion that individuals who develop ASD might have an imbalance between paternally and maternally imprinted genes. More specifically, they have an overabundance of paternally active genes and these genes proceed to enable greater resource extraction from the mother during foetal growth and during infant life, leading to the empirically observed higher growth rates and greater weights and lengths compared to children who go through typical neurodevelopment.

The objective of this current study is to compare children who are already diagnosed with ASD to typically developing controls in regard to the anthropometric measurements, in the context of a putative genomic imprinting imbalance.

## METHODS

Our study included two groups of male children, with ages between 4 and 8 years old: 30 individuals in the ASD group and 33 individuals and the control group. Children presenting in the paediatric out-patient service between 2014-2018 were included in the study as well as children (both those with typical neurodevelopment and those with a formal ASD diagnosis) from local community kindergartens, primary schools and patient-oriented community services.

Informed consent was obtained from parents or guardians during the initial consultation and prior to including measurements in the dataset. The study design was approved by the local Ethics Committee of the Cluj-Napoca Emergency Clinical Children’s Hospital.

Height, weight, head circumference and thoracic circumference were measured. All measurements were evaluated for the normality of their distribution by the Shapiro-Wilk test and subsequent comparison of the means was made using Welch’s two-sample t-test. Significance level was placed at p<0.01 due to multiple comparisons of the same group.

## RESULTS

No difference in age between groups: ASD group 64.9 months versus Control group 67.6 months (p=0.49).

Children with ASD are no different in size than controls in this ages group (see Table 1).

**Table 1.**
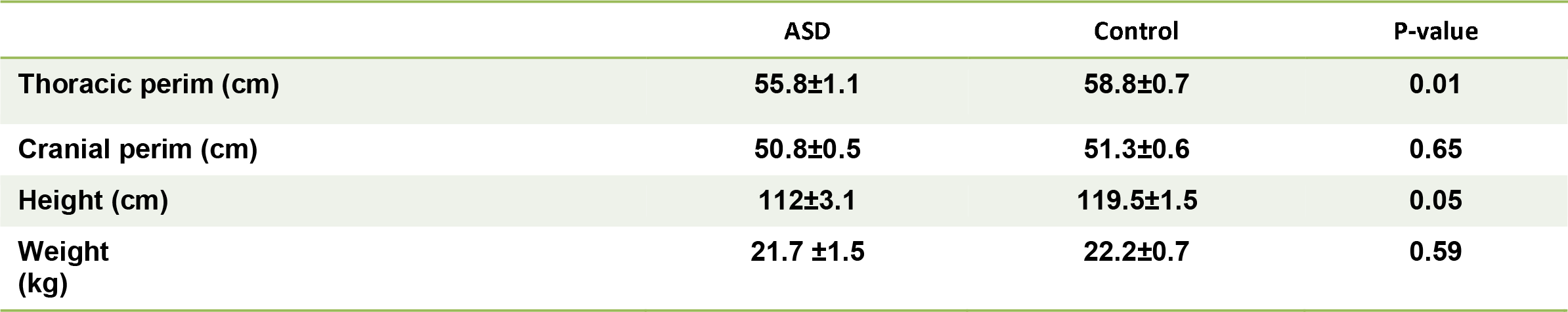
Results are presented as mean ± standard deviation of the mean.

## DISCUSSION

As matters stand right now, there seem to be **no differences between children with ASD and neurotypical controls** regarding somatic growth by age 4-8.

Some somatic measurements in this study population show a trend towards being greater in neurotypical controls.

These results bring an interesting dynamic to the discussion regarding the role of imprinted genes in the development of ASD.

As previously explained^3^, **imprinted genes can be expected to change their expression pattern in order to provide relative benefits to the parent of origin**^8^. Paternally active genes would increase resource extraction from the mother while the infant is highly dependent on the mother (during development and breastfeeding). **This would explain the higher birth weight and length and early overgrowth**. However, **after weaning, relative paternal contribution ought to increase**, thus one would expect paternally active genes to start changing the infant’s behavior, so as to make the child less demanding of resources. Thus, they might start lagging behind in somatic growth as overall resource extraction capacity decreases. **This would explain the data presented here, that shows children with ASD being no different from controls**.

**The underlying mechanism of autism spectrum disorder pathogenesis described herein as the genomic, hereditary approach, coincides well with several other proposed mechanisms and clinical observation made in relation to children with ASD, at different types of approaches**.

**First - the phenomenological, clinical approach. Re**gression has been a feature of clinical descriptions of autism. Though it lacks a clear consensus among the professional community as to its prevalence and indeed, its neurobiological significance^10^, clinical features of regression (loss of acquired developmental and social skills), are observed in certain subsets of patients later diagnosed with ASD. Since these clinical features only become apparent as development progresses, one must account for the underlying biology of this essentially phenomenological manifestation of autism.

In relation to the notion proposed in this paper, that imprinting imbalances contribute to the development of ASD, the phenomenon of regression should be interpreted as a continuous process, that only becomes evident at certain ages when particular milestones are observed to be missing or diminishing. This is consistent with the idea that, as the phenotypical effects of the genetic imprint shifts, it affects neurodevelopment and it might be possible for this shift to outwardly manifest in the behavioral changes described as regression.

**Second - the neuroanatomical approach. A**s these genomic imprinting imbalances would only constitute the instructions of heredity for different biological processes, the question arises of what might these processes be. There is ample evidence that, at the neuroanatomical level, patients who develop ASD suffer from defects of the normal activity of synaptic pruning^11^. As synaptic pruning unfolds throughout a child’s development, genetic imprinting imbalances could presumably affect and shift these processes into low or high atypical pruning thresholds. Excessive pruning might damage otherwise hitherto typically developed neural circuitry^11^ or insufficient pruning might leave extraneous, inefficient synaptic connectivity in local circuitry^12^.

**Additional circumstantial evidence** supporting an inter-relation of these different approaches come from the fact that a formal and medically well-grounded ASD diagnosis is mostly made in kids who older than 3 years of age ^13^. Considering that, on average, the second year of the child’s life marks a slow transition from almost complete dependence on the mother to a more dynamic interaction with care-givers and subsequent relative increase of paternal input into child-rearing, it would be expected that the paternally-biased imprinted genome would gradually shift the offspring’s behavior from extracting more and more resources from the mother to not being a significant burden on father.

Consequently, it is interesting to note that many phenomenological features of regression, as anecdotally presented by care-givers in the clinic also take part during the second year of life ^14^. Moreover, synaptic pruning starts during infancy and involves motor and sensory areas but synaptic pruning continues well into middle childhood and adolescence with temporal and prefrontal^11^. This is consistent with reports of regression involving loss of motor skills or reduction in basic speech sounds, reported on average before the child’s second birthday^14^ and might constitute the basis of deepening social deficits seen later on that constitute the diagnostic criteria of ASD. Thus, the progression of behavioral features associated with autism during middle childhood and later on are consistent with the notion that a more imbalanced imprint, with a relative overabundance of paternally active genes, drives neuroanatomical changes, potentially by means of synaptic pruning.

## CONCLUSIONS

Given the fact that children with ASD seem to get a head start in growth, as published in the scientific literature to date^5,6,7,8,^ the lack of differences found in our 4-8 years old group indicates that **children with ASD might actually fall behind in somatic growth, or at least stagnate**.

There is also evidence placing the onset of observable behavioral changes (either as loss of previously acquired motor and linguistic skills or disturbances of social development) during the period when paternally active genes are expected to start shifting behavior from “resource extraction from the mother” to “lessening the resource burden demanded from the father”.

Relative overabundance of paternally active genes could explain these results.

Stronger proof for these hypotheses could come from following a large cohort from birth all the way up to early teenage years – as in Byars et al^7^.

Clinicians should bear in mind the fact that children with ASD might fall behind in somatic growth.

## Data Availability

Data availability: upon requet to author.

